# Efficacy of a patient isolation hood in reducing exposure to airborne infectious virus in a simulated healthcare setting

**DOI:** 10.1101/2022.07.24.22277784

**Authors:** Leo Yi Yang Lee, Shane A Landry, Milan Jamriska, Dinesh Subedi, Simon A Joosten, Jeremy J Barr, Reece Brown, Kevin Kevin, Robyn Schofield, Jason Monty, Kanta Subbarao, Forbes McGain

**Author notes:** **Corresponding author:** Dr. Forbes McGain (footnotes), **Corresponding author contact details:** Dr. Forbes McGain, Contact address: Sunshine Hospital, Furlong Road, St Albans, VIC 3021, Australia.

## Abstract

**Background:** Healthcare workers treating patients with SARS-CoV-2 are at risk of infection from patient-emitted virus-laden aerosols. We quantified the reduction of airborne infectious virus in a simulated hospital room when a ventilated patient isolation (McMonty) hood was in use.

**Methods:** We nebulised 10^9^ plaque forming units (PFU) of bacteriophage PhiX174 virus into a 35.1m^3^ room with a hood active or inactive. The airborne concentration of infectious virus was measured by BioSpot-VIVAS and settle plates using plaque assay quantification on the bacterial host *Escherichia coli C*. The particle number concentration (PNC) was monitored continuously using an optical particle sizer.

**Results:** Median airborne viral concentration in the room reached 1.41 × 10^5^ PFU.m^-3^ with the hood inactive. Using the active hood as source containment reduced infectious virus concentration by 374-fold in air samples. This was associated with a 109-fold reduction in total airborne particle number escape rate. The deposition of infectious virus on the surface of settle plates was reduced by 87-fold.

**Conclusions:** The isolation hood significantly reduced airborne infectious virus exposure in a simulated hospital room. Our findings support the use of the hood to limit exposure of healthcare workers to airborne virus in clinical environments.

**Lay summary:** COVID-19 patients exhale aerosol particles which can potentially carry infectious viruses into the hospital environment, putting healthcare workers at risk of infection. This risk can be reduced by proper use of personal protective equipment (PPE) to protect workers from virus exposure. More effective strategies, however, aim to provide source control, reducing the amount of virus-contaminated air that is exhaled into the hospital room.

The McMonty isolation hood has been developed to trap and decontaminate the air around an infected patient. We tested the efficacy of the hood using a live virus model to mimic a COVID-19 patient in a hospital room. Using the McMonty hood reduced the amount of exhaled air particles in the room by over 109-times. In our tests, people working in the room were exposed to 374-times less infectious virus in the air, and room surfaces were 87-times less contaminated. Our study supports using devices like the McMonty hood in combination with PPE to keep healthcare workers safe from virus exposure at work.

## Background

The treatment of patients with COVID-19 has led to considerable risk of infection in healthcare workers (HCW) with associated morbidity and mortality [1, 2]. SARS-CoV-2 virus can be spread by fomites, droplets and aerosol particles [2, 3]. The U.S. National Institute for Occupational Safety and Health (NIOSH) hierarchy of hazard controls ranks engineering approaches to isolate people from the source of risk above personal protective equipment (PPE) as methods to reduce exposure risks [4]. Nevertheless, much focus has been placed upon correct PPE use by HCW (e.g. N95 mask and barrier gown wearing) in lieu of improved engineering controls to reduce cross-infection of SARS-CoV-2 to HCWs and other patients [3].

Hospital environmental engineering controls primarily rely upon negative pressure rooms (NPRs) to limit the spread of airborne viruses outside of designated areas. However, NPRs are a scarce resource, may not fully contain SARS-CoV-2, and importantly, may not entirely protect HCWs within the NPR [5]. Another engineering control that is gaining traction is the use of portable air cleaners to enhance clearance of contaminated air around HCWs, which can be used in concert with existing measures [6, 7]. It would be ideal to directly control the emission source of infectious aerosols to reduce the transmission risk.However, there are limited methods to contain the respiratory aerosols emitted by infectious patients, e.g. placing an N95 mask on the patient, which cannot be done without compromising their care. Prior to the COVID-19 pandemic, personal isolation hoods were rarely used for respiratory virus outbreaks [8], although interest has recently increased [9, 10]. Several members of our research group have developed a personal isolation hood (McMonty hood) [11] and examined its utility in reducing exposure to physical aerosols by at least 98% in a clinical test environment [12]. Subsequently this isolation hood has been adopted by many hospitals in metropolitan and regional Australia.

The efficacy of the isolation hood in reducing airborne viral load has not been validated, and the claimed efficacy was simply derived from the reduction in the physical aerosol counts. Ideally we would have tested the isolation hood’s effectiveness in containing SARS-CoV-2 directly in a hospital. However, such a clinical approach: (i) would have exposed research staff to patients with COVID-19; (ii) would not be controlled (i.e. there would be considerable variability in aerosol emission between different patients), and; (iii) would be less likely to ‘stress’ the isolation hood to prolonged worst-case scenarios with a maximum number of viruses for extended durations.

The bacteriophage PhiX174 (family *Microviridae*) is a small (25nm, approx. ¼ SARS-CoV-2’s size) [13], non-enveloped, bacteriophage with a linear ssDNA genome that is harmless to humans and is routinely used as a surrogate pathogen for the study of airborne viral transmission [14-16]. Landry et al. [17] recently quantified viable airborne PhiX174 virus propagated from a positive airway pressure circuit leak. Nebulised viral aerosols were successfully contained by a makeshift plastic hood cover and a commercial HEPA filter with a fan [17]. We aimed to test how effectively the McMonty patient isolation hood could actively contain an airborne virus emission source by nebulisation of the surrogate virus PhiX174, simulating the hood’s ability to limit the risk of infectious aerosol exposure to HCWs in clinical settings [18].

## Methods

Ethics approval for this laboratory-based study was deemed not required by Monash University’s School of Biological Sciences Ethics Manager.

### Bacteriophage propagation and quantification

Bacteriophage PhiX174 was propagated on its bacterial host *Escherichia coli C* (ATCC13706) grown in lysogeny broth (LB). Viral product was purified from lysate using the Phage-on-Tap protocol [19] and resuspended in 1X phosphate-buffered saline (PBS, Omnipur,® Merck, Gibbstown, NJ, USA). The viable concentration of PhiX174 virus stocks was quantitated by plaque assay using the soft agar overlay method [19]. Viable counts of PhiX174 were expressed as plaque forming units per millilitre of suspension (PFU/mL).

### Patient isolation hood

The McMonty isolation hood consists of a mobile steel frame, a plastic canopy, and an extraction fan equipped with a standard high efficiency particulate air (HEPA) H13 filter (rated to 99.95% clearance of 0.3 µm particles). The plastic barrier opens out to form a hood with 1.3m^3^ internal volume, enclosing the patient’s torso from the waist up (non-airtight seal) (Figure 1). The extraction fan (Westaflex, Melbourne, Australia) is mounted behind and above the patient’s head. It draws the air from around the patient, passing it through a HEPA filter (Techtronic Industries, Hong Kong, China) before recirculating clean air to the surroundings (clean air delivery rate: CADR, at 144m^3^/h) [12].

**Figure 1.**
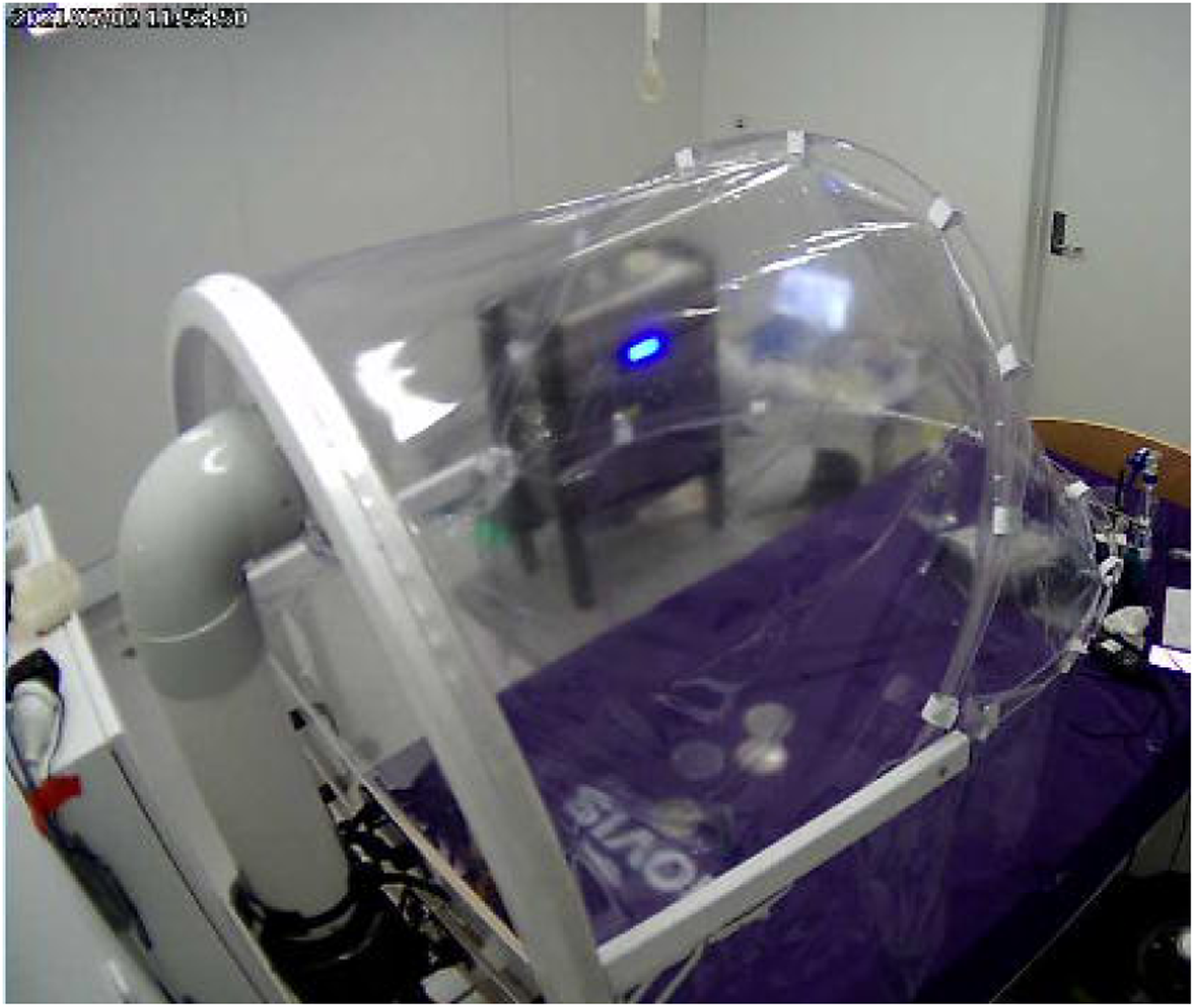
McMonty personal isolation hood in active configuration. Hood deployed in simulated hospital room in experiment conditions. VIVAS instrument and settle plates visibly arranged around mock patient bed. Photo credit: Shane Landry (from experiment monitoring camera).

### Simulation of airborne virus containment using the McMonty hood in a hospital room

The McMonty isolation hood was tested in a simulated hospital room that contained a single bed with the hood positioned to isolate a simulated patient. PhiX174 bacteriophage was aerosolised into the sealed room (4m x 3.5m x 2.4m, 35m^3^ internal volume) to mimic the shedding of airborne virus. The virus emission source was simulated using a nebuliser (Pari-PEP®, PARI Respiratory Equipment, VA, USA) placed where the head of the patient would rest (20cm above surface of bed, see Figure 2 location N) with the outlet facing upwards. The Pari-PEP device produces aerosol particles with unimodal polydisperse size distribution with the mass median diameter of 3.42 ± 0.15 μm [20]. The room ventilation ports were covered to avoid viral egress (i.e., no active HVAC). Sampling devices were arranged around the room to detect the spread of airborne particles laden with PhiX174 (Figure 2). Room temperature and relative humidity during all experiments was monitored continuously and was in the range of 21.6-27°C (mean 24.7°C) and 46-67% (mean 53.1%), respectively.

**Figure 2.**
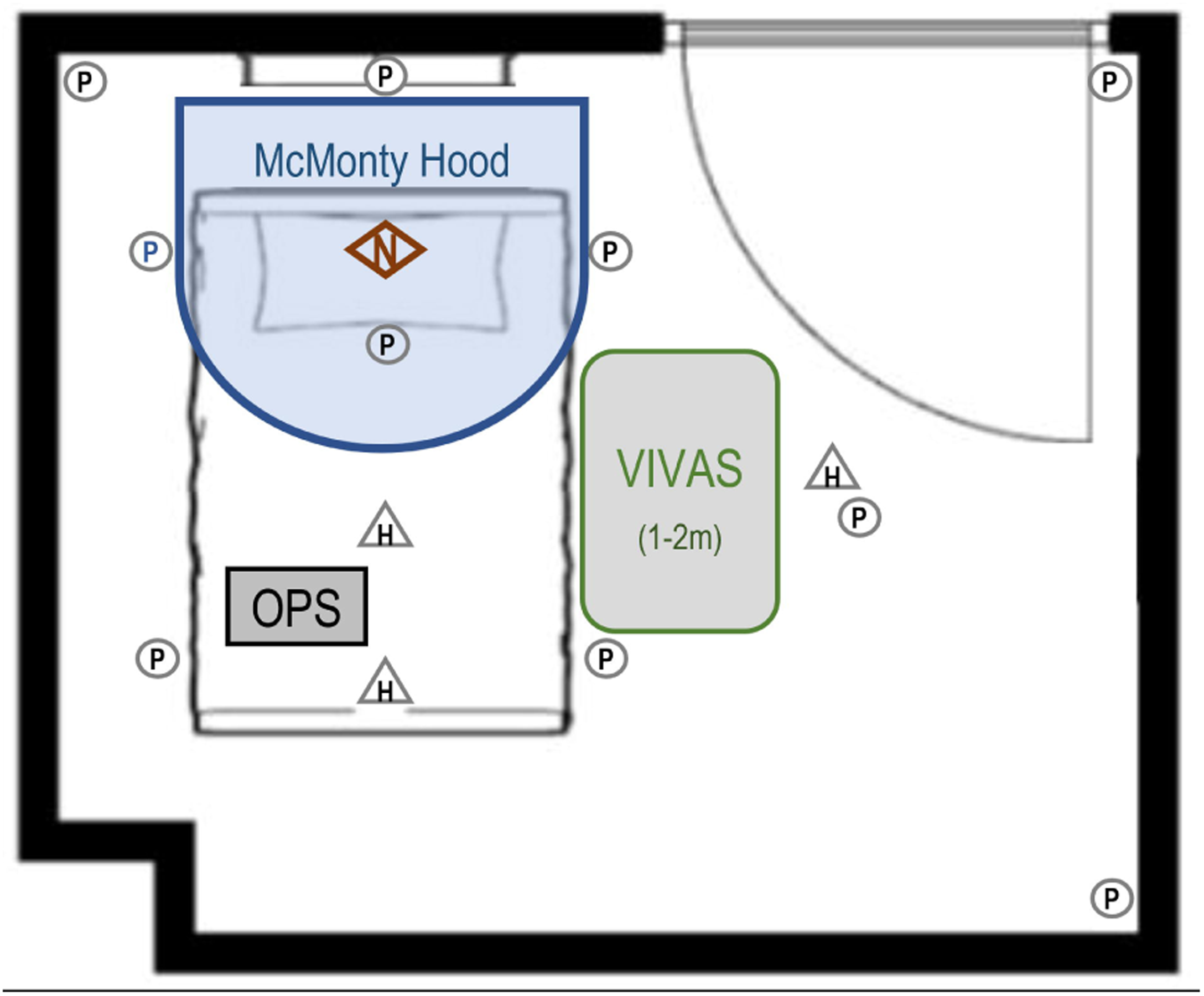
Locations of settle plates, the VIVAS machine and OPS in the simulation room. All experiments were performed in a clinical room with dimensions 4.0 × 3.25 × 2.7 m (volume = 35.1 m^3^) containing a bed and the McMonty hood. Ten settle plates (grey circles) and three hanging plates (grey triangles) were hung at head height perpendicular to the floor. The nebuliser (N, orange diamond) was positioned at the head of the bed, with the exit point facing vertically upwards. Air sampling was performed using a BioSpot VIVAS positioned at the bedside 1-2 m from the nebuliser. Particle concentration as assessed by optical particle sizer (OPS) posited in the centre of the bed, outside the hood. Virus and aerosol measurements were taken when the nebuliser was actively contained by the hood (i.e., hood physically isolated nebuliser from instruments), compared to when no containment was used (i.e., instruments were exposed to nebuliser).

Aerosol generation experiments were conducted four times each day over three independent days during July-September 2021. For each experiment, 10mL of PhiX174 virus suspension was completely nebulised (airflow at 9 litres per minute, lpm) for a 40 min period. In parallel, all virus and aerosol sampling devices were exposed to the air in the room for the duration. Experiment conditions were: (i) McMonty hood providing active source containment, where the plastic barrier was deployed with the fan running to enclose the nebuliser; or (ii) hood inactive (no containment), where the fan and plastic barrier were not deployed and the nebuliser was exposed to the room. Following each nebulisation period, the room air was purged of aerosols by running a portable air purifier (IQAir HealthPro 250, Goldach, Switzerland) for 30 mins at a CADR of 470 m^3^/h before beginning the next experiment.

### Detection of airborne infectious PhiX174

We employed the passive settle plate detection method for nebulised PhiX174 established previously by Landry et al. [17] Thirteen settle plates were positioned in the room and exposed during each simulation (Figure 2) to detect the deposition of airborne particles laden with infectious virus.

In parallel, the BioSpot-VIVAS 300-P (VIVAS; Aerosol Devices, Fort Collins, CO, USA) was used to actively sample particles from air in the room at 8 lpm. The VIVAS was positioned adjacent to the hospital bed and its air sample intake was located 1-2m away from the nebuliser (Figure 2). Each air sample was collected as a condensed fluid into a petri dish (35mm diameter) containing 3 mL of sterile PBS, which was stored at 4°C until analysis. The infectious titre of PhiX174 in VIVAS sample fluid was determined by plaque assay on soft agar overlay plates of *E. coli C* as described above. VIVAS detection of airborne virus was expressed as PFU per cubic metre of indoor air collected during the 40 min nebulisation period (PFU.m^-3^). The VIVAS inlet and sampling lines were decontaminated (Supplementary Figure S2) by flushing with 70% ethanol then distilled water prior to the next sample.

### Aerosol monitoring instrumentation

Airborne particle number distribution and concentration were monitored during each experiment using an optical particle sizer (OPS, TSI Model 3300), which allows detection and size classification of aerosol particles within a 0.3–10µm diameter range. McMonty hood performance was assessed using total particle number concentration (PNC). The instrument was placed outside of the hood at the foot of the patient bed with sampling inlet 0.2m above the bed (Figure 2) and logged measurements at 10 second average intervals. The OPS was present for two out of three independent experimental days (Supplementary Materials).

### Data analysis

Infectious phage samples from each experiment were categorised based on their relative exposure to the nebuliser with the McMonty hood active or inactive. Data from all simulation samples were treated as experimental replicates and pooled according to containment condition. Settle plates counts exceeding the limit of detection were included as 300 PFU. The untransformed PFU counts (settle plate) or PFU.m^-3^ (VIVAS) were compared between each containment condition by the Wilcoxon rank sum test (R version 4.1.1, The R Foundation for Statistical Computing, Vienna; package ‘rstatix’). P values <0.05 were considered statistically significant. Wilcoxon effect size (r) was calculated as Z score/√(sample size). For both settle plate and VIVAS data, we compared the fold-change in median values for McMonty hood active versus inactive datasets to express the reduction in bacteriophage resulting from the use of active source containment.

We defined a theoretical model to describe the PNC inside the room throughout each experiment (Supplementary Materials). The model explains rate of change in PNC over time (per minute) through a combination of (i) the number of viral aerosol particles escaping the hood; and (ii) the number of viral aerosol particles in the room lost through deposition. Experimental PNC data measured by the OPS was fitted to this model to calculate the number of particles escaping into the room when the McMonty hood was active versus inactive. These values were used to calculate the effective filtration efficiency, expressed as the percentage (%) of particles the hood prevents from escaping into the room.

## Results

### Patient isolation using the McMonty hood reduces concentration of airborne infectious virus

Figure 3 shows the airborne concentration of infectious PhiX174 in the room using an active McMonty hood compared to an inactive control condition (no containment). With active containment of the emission source, the median concentration of viable virus in air samples was 3.77 × 10^2^ PFU.m^-3^ (n=6, IQR=262-1,277) compared to 1.41 × 10^5^ PFU.m^-3^ (n=7, IQR=33,750-182,500) without containment. This equates to a significant 374-fold reduction in airborne infectious virus contamination (W=0, p<0.005, r=0.83).

**Figure 3.**
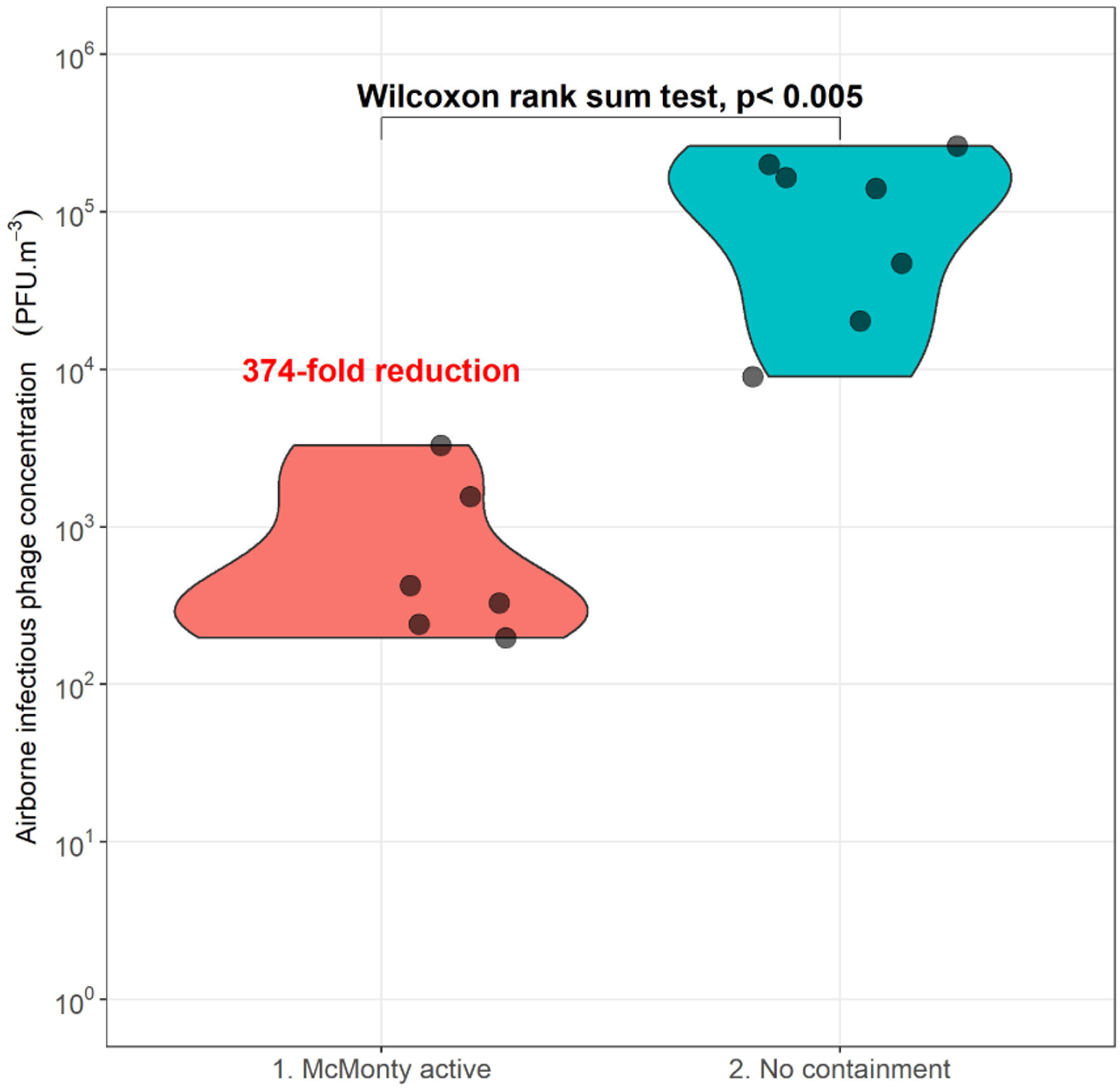
Violin-plot of viable airborne phage concentration (PFU.m^-3^) detected by Bio-Spot VIVAS (y-axis) in each containment condition (x-axis). Distribution of individual measurements (black points) of bacteriophage plaque forming units per m^3^ of air (PFU.m^-3^) when the McMonty hood was inactive during the sampling period (blue fill) compared to measurements taken when active hood source containment was used (red fill). Annotated 374-fold reduction of median airborne virus concentration when McMonty hood was active. Groups were compared (untransformed values) by Wilcoxon rank sum test.

### Effect of the Active McMonty hood upon containment of airborne particles

Airborne particle number concentrations (PNCs) for McMonty active and inactive no containment conditions are shown in Figure 4. Both datasets were fitted to a model (Supplementary Materials) to quantify the rate of viral aerosol escaping into the simulation room in each case. One room simulation with the McMonty hood active was excluded from analysis due to data inconsistencies (Supplementary Figure S1).

**Figure 4.**
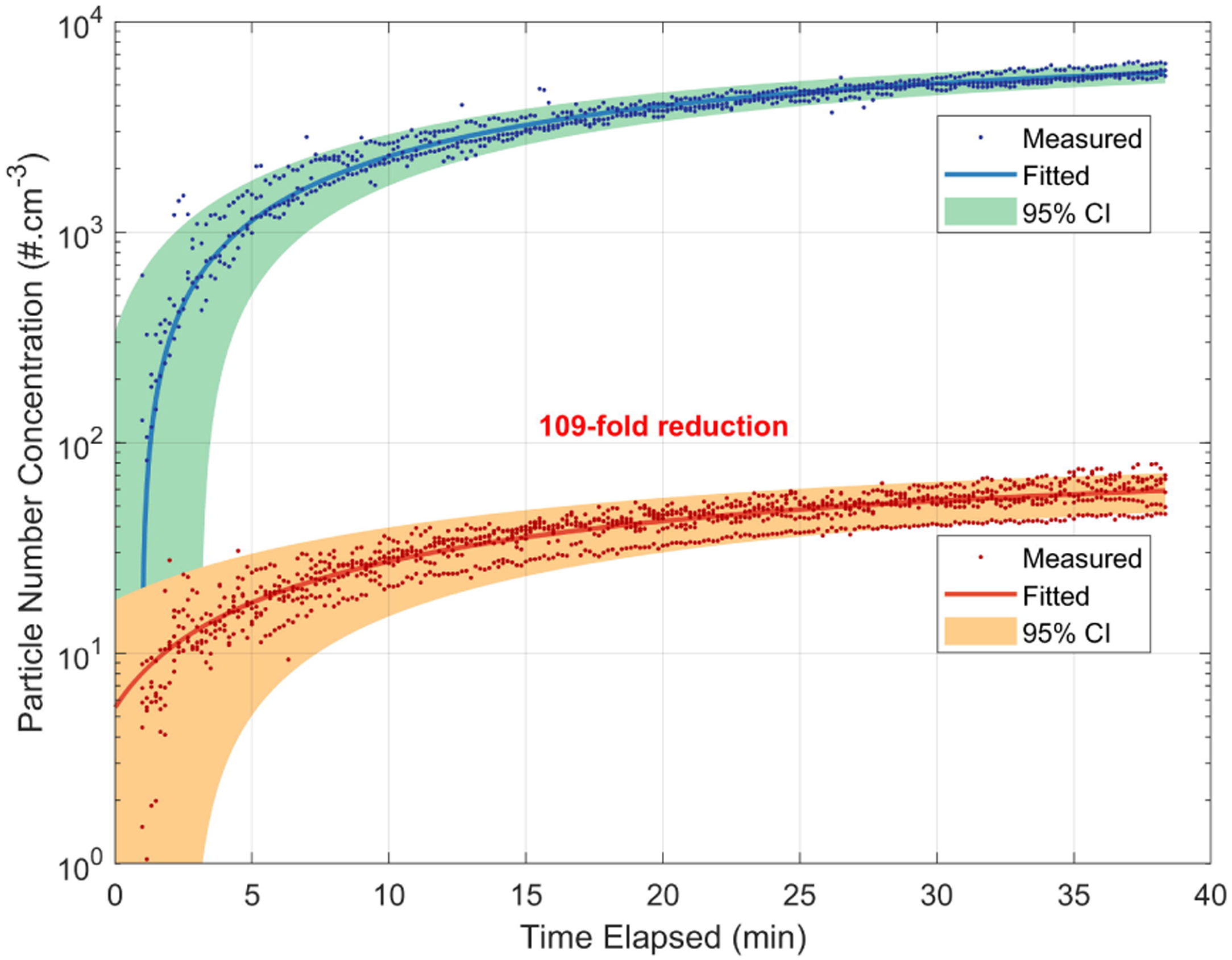
PNC data from OPS measurements taken during airborne phage experiments. Data grouped based on containment type and fitted to theoretical model with 95% confidence interval. Labelled fold-reduction of particle number escape rate when McMonty hood is active (data points in red) compared to inactive (data points in blue).

The combined PNC data for each condition show clear common trends and limited variability, indicating good reproducibility between each experiment. Furthermore, the fitted model shows good agreement with both datasets, with adjusted r^2^ values of 0.86 when the McMonty Hood is active and 0.96 when inactive.

A clear 2 orders of magnitude decrease in PNC was observed within the room when the McMonty hood is active (Figure 4). This large reduction was also reflected in the modelled escape rates (Table 1) with a 109 ± 5 fold reduction in the rate of viral aerosol escape into the room when the hood is active. Effective filtration efficiency of the hood (calculations in supplement) indicated that the active McMonty hood successfully removed 99.1 ± 0.1 % of viral aerosol released. Comparing total airborne particle (via OPS) and infectious airborne virus (via VIVAS) measurements shows that active McMonty hood containment mitigates >99% of respiratory exposure in our simulated hospital room.

**Table 1.** Summary of physical and infectious aerosol results from OPS and VIVAS. Fitted values of viral aerosol escape rates (± mean standard deviations) with the McMonty hood active or inactive are presented, with the calculated fold reduction and effective filtration efficiency. Median virus concentration (± median absolute deviation) measurements are provided for VIVAS virological data; corresponding fold reduction and percentage reductions are calculated from the median values.

| Physical Aerosol Analysis | McMonty Active Escape Rate ($\log_{10}\#\cdot\text{min}^{-1}$) | Inactive (No Containment) Escape Rate ($\log_{10}\#\cdot\text{min}^{-1}$) | Fold Reduction | Effective Filtration Efficiency (%) |
| --- | --- | --- | --- | --- |
| | 7.99 ± 6.60 | 10.02 ± 8.48 | 109 ± 5 | 99.1 ± 0.1 |
| Virological Aerosol Analysis | McMonty Active Viral Conc. (median) ($\log_{10}\text{PFU}\cdot\text{m}^{-3}$) | Inactive Viral Conc. (median) ($\log_{10}\text{PFU}\cdot\text{m}^{-3}$) | Fold Reduction | Percentage Reduction (%) |
| | 2.58 ± 2.36 | 5.15 ± 5.14 | 374 | 99.7 |

### Infectious virus counts from airborne particles which deposited onto the surface of settle plates

Infectious virus counts on settle plates were reduced with active source containment (<11 PFU), and 33/108 plates did not detect any viable phage. Active isolation hood containment significantly reduced the median settle plate count (median=2, n=108 IQR=0-4) by 87-fold (W=91.5, p<0.001, r=0.775) compared to plates without source containment (median=174, n=48, IQR=94.5-300) (Figure 5).

**Figure 5.**
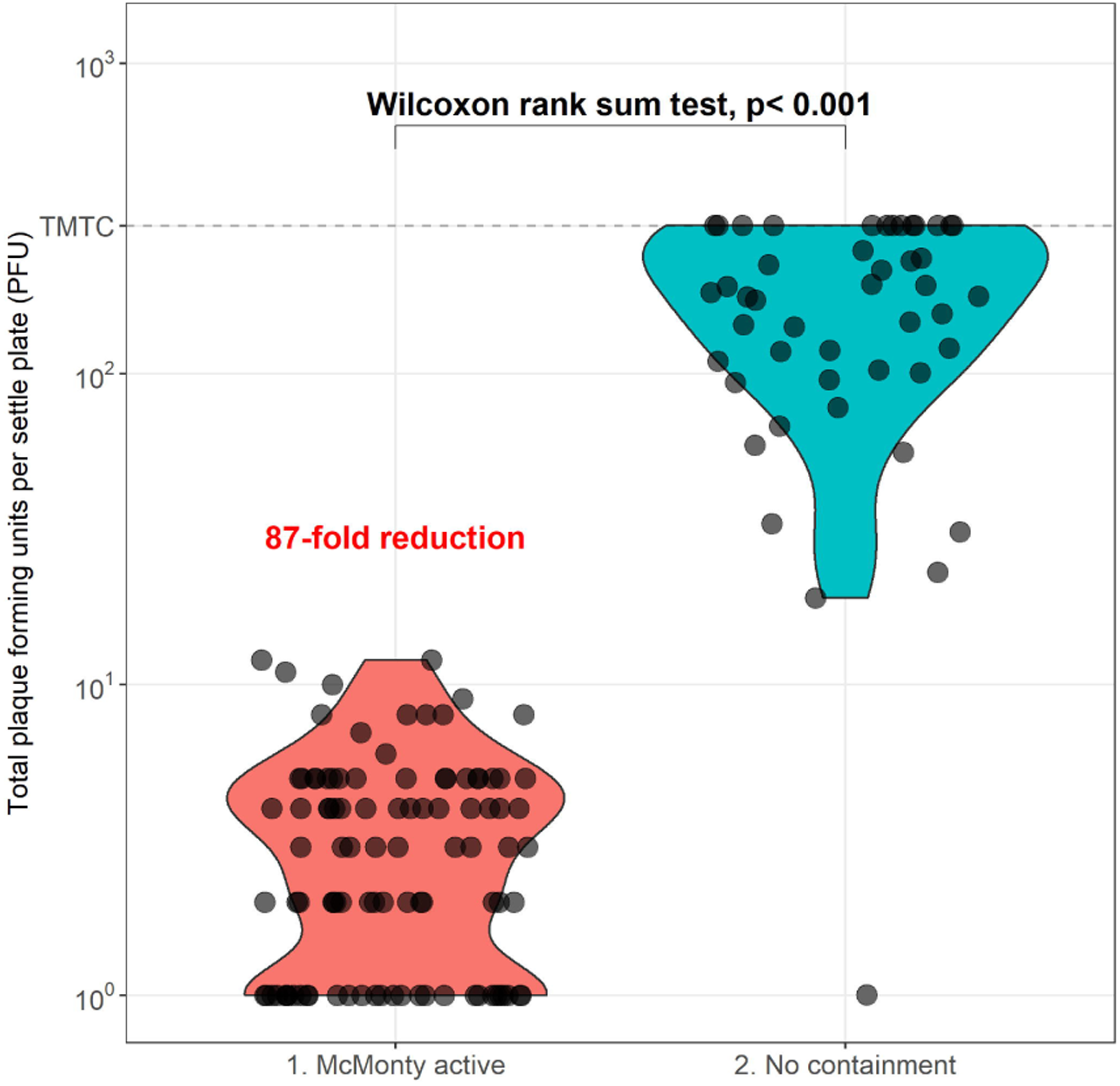
Violin-plot of total viable phage count (PFU) detected on settle plates (y-axis) in each containment condition (x-axis). Distribution of individual plaque forming unit (PFU) counts (black points) for each settle plate when no containment (inactive hood) was used during the sampling period (blue fill) compared to counts reached when active McMonty hood containment was used (red fill). Counts transformed by +1 PFU to visualise 0 PFU plates on the log_10_ scale (on graph only). Plates with plaque counts exceeding the saturation point on each plate were given the value of 300 for statistical tests (threshold labelled as too many to count; TMTC). Annotated 87-fold reduction of median settle plate count when McMonty hood is active. Groups were compared (untransformed values) by Wilcoxon rank sum test, p < 0.05 indicates statistically significant difference. Outliers were excluded from violin-plot visualisations but are visible in the scatterplot.

## Discussion

In a simulated patient room, we nebulised a high titre bacteriophage (PhiX174) suspension, comparing infectious phage numbers and particle number concentrations (PNCs) when a McMonty isolation hood was active/inactive. Measurements of virus-laden aerosols using the Bio-Spot VIVAS showed a 374-fold reduction in infectious virus concentration, and a 109-fold reduction in particle escape rates when the isolation hood was active. That is, viral aerosol spread was reduced by >99% when the isolation hood was active, consistent with our prior study using non-infectious aerosols [12]. There was also an 87-fold reduction in viable virus deposited onto bacterial settle plates when the hood was active, indicating a reduced risk of contamination of surfaces.

Nielsen et al. [8] in 2009 discussed the utility of personalized ventilation as part of a suite of indoor controls of airborne infectious diseases. Johnson et al. (2009) found that effective isolation was possible using feasible, low-technology, low-cost structures readily constructed within hospitals in emergency situations [21]. Since the COVID-19 pandemic several groups have focused upon aerosol containment at the point of emission with personal isolation units, including ventilated headboards [9], and similar stationary devices [10] attached to building ventilation units. Nishimura et al. atomised influenza virus inside their Barrihood isolation fan/filter unit, finding that none escaped [22]. Using a bacteriophage model similar to our study, Landry et al. (2022b) showed that a makeshift isolation hood greatly augments the protection conferred by standard hospital PPE by limiting skin exposure to viral aerosols [23]. Despite the success of these isolation hood concepts in experimental settings, such devices have not been in widespread use by HCWs during the COVID-19 pandemic.

Concessions to human comfort requirements must be addressed for the adoption of isolation hoods in clinical settings. Correlating well with our previous report using non-infectious aerosols [12], we found that the McMonty hood achieved a 99.1% effective filtration efficiency by aerosol particle counts. This was lower than the theoretical 99.95% filtration efficiency of its H13 HEPA unit [12], indicating that the small fraction of viral aerosol escaping the hood was likely diffusing underneath the hood skirting rather than penetrating through the exhaust filter. This trade-off aims to improve patient comfort and usability by maintaining lower (<0.5 m/s) airflow rates near the patient [24], while cognisant that lower air flow rates reduce the efficacy of clearing infectious respiratory agents.

Our study has some limitations. First, while the bacteriophage PhiX174 is a safe surrogate for airborne viral spread, it is unenveloped, unlike coronaviruses and influenza viruses, which may affect its environmental stability [17]. Second, we analysed airborne and settle plate viral contamination in a laboratory room that simulates a functionally uniform aerial viral distribution, in the absence of either active HVAC or air mixing. Similarly, the concentrated viral inoculum nebulised into our simulation room was several orders of magnitude higher than previously reported levels of airborne infectious virus shed by COVID-19 patients [25-27]. The detection of approx. 10^3^ PFU.m^-3^ airborne phage escaping hood containment is likely a result of this amplified viral challenge. Combined, these environmental and viral load factors produce an extreme test scenario. Finally, we used a nebuliser which produces a relatively narrow range of particle sizes (aerosol mass median diameter 3.42 ± 0.15μm) to produce virus-laden aerosols. Human generated aerosols are of similar mean size [28], though with a broader size distribution, particularly for large, visible droplets [29].

We employed two methods to quantitate infectious airborne virus levels to address different types of exposure risk. Bacteriophage counts from the VIVAS were approximately 100-fold higher than floor settle plates, as the device actively pumps large air volumes to capture suspended infectious aerosols into a liquid sample. In contrast, the bacterial plates relied on passive deposition of aerosols onto <1% of the floor surface area, thus high viral loads/concentrations were required to account for this difference in sensitivity [17]. The VIVAS may have further reduced the sensitivity of the plate counts by competition as it was drawing in virus-laden air away from the plates. These sampling techniques can be correlated to different risk factors in a hospital environment involving airborne viral contamination. The respiratory exposure risk of medical staff in virus-infected patient rooms can be approximated by the VIVAS air sampling rate of 8 lpm, which is similar to human minute ventilation rates (7 lpm) [30]. The deposition of infectious virus aerosols onto the surface of settle plates can similarly relate to the potential risk of fomite generation on contaminated room surfaces, although our experimental setting lacked realistic airflow by HVAC. In our test system, the McMonty hood substantially reduced viral exposure by both these metrics.

We have shown that a personal isolation hood can effectively reduce viral and aerosol escape by >99% in a simulated indoor healthcare setting. Along with our recent clinical study of ease-of-use [31], these findings support the clinical use of the hood to limit HCW exposure to airborne virus. Complementary efforts to clean indoor air environments with HEPA filtered air cleaners in hospitals [6, 7] and beyond are also useful to prevent respiratory infections. Undertaking a randomised, controlled clinical trial of the efficacy of isolation hoods (or of air cleaners) in preventing HCW SARS-CoV-2 infections is likely to be overwhelmingly challenging due to the large sample size required.

## Data Availability

All data produced in the present study are available upon reasonable request to the authors.

## Footnotes

### Authors contributions

LYYL, SAL, SJ, MJ, JB, KS and FM contributed to study concept and design; LYYL, SAL, SJ, MJ, DS, JB, and FM contributed to acquisition of the data, LYYL, DS, RB, MJ, SAL, SJ and FM contributed to data analysis and interpretation; LYYL, FM, RB, MJ, DS and JB contributed to the initial drafting of the manuscript; all authors contributed to critical revision of the report.

## References

1. Bandyopadhyay S, Baticulon RE, Kadhum M, et al. Infection and mortality of healthcare workers worldwide from COVID-19: a systematic review. BMJ global health 2020; 5:e003097.

2. World Health Organization. Transmission of SARS-CoV-2: implications for infection prevention precautions. Available at: https://www.who.int/news-room/commentaries/detail/transmission-of-sars-cov-2-implications-for-infection-prevention-precautions.

3. Morawska L, Cao J. Airborne transmission of SARS-CoV-2: the world should face the reality. Environment International 2020; 2020 Apr 10;139:105730. doi: 10.1016/j.envint.2020.105730:105730.

4. (NIOSH) TNIfOSaH. Hierarchy of Controls. Available at: https://www.cdc.gov/niosh/topics/hierarchy/default.html.

5. Santarpia JL, Rivera DN, Herrera VL, et al. Aerosol and surface contamination of SARS-CoV-2 observed in quarantine and isolation care. Scientific reports 2020; 10:1–8.

6. Buising KL, Schofield R, Irving L, et al. Use of portable air cleaners to reduce aerosol transmission on a hospital COVID-19 ward. medRxiv 2021.

7. Landry SA, Subedi D, Barr JJ, et al. Fit-tested N95 masks combined with portable HEPA filtration can protect against high aerosolized viral loads over prolonged periods at close range. The Journal of Infectious Diseases 2022a.

8. Nielsen PV. Control of airborne infectious diseases in ventilated spaces. Journal of the Royal Society Interface 2009; 6:S747–S55.

9. Centers for Disease Control and Prevention. NIOSH Ventilated Headboard Provides Solution to Patient Isolation During an Epidemic. Available at: https://blogs.cdc.gov/niosh-science-blog/2020/04/14/ventilated-headboard/?deliveryName=USCDC-170-DM25875.

10. Adir Y, Segol O, Kompaniets D, et al. Covid19: minimising risk to healthcare workers during aerosol producing respiratory therapy using an innovative constant flow canopy. European Respiratory Journal 2020; 55:2001017.

11. Medihood. The McMonty Medihood. Available at: https://medihood.com.au/.

12. McGain F, Humphries RS, Lee JH, et al. Aerosol generation related to respiratory interventions and the effectiveness of a personal ventilation hood. Crit Care Resusc 2020.

13. Bar-On YM, Flamholz A, Phillips R, Milo R. SARS-CoV-2 (COVID-19) by the numbers. Elife 2020; 9:e57309.

14. Verreault D, Moineau S, Duchaine C. Methods for sampling of airborne viruses. Microbiology and molecular biology reviews 2008; 72:413–44.

15. Turgeon N, Toulouse M-J, Martel B, Moineau S, Duchaine C. Comparison of five bacteriophages as models for viral aerosol studies. Applied and environmental microbiology 2014; 80:4242–50.

16. Kumar S, Nyodu R, Maurya VK, Saxena SK. Morphology, Genome Organization, Replication, and Pathogenesis of Severe Acute Respiratory Syndrome Coronavirus 2 (SARS-CoV-2). Coronavirus Disease 2019 (COVID-19) 2020:23–31.

17. Landry SA, Barr JJ, MacDonald MI, et al. Viable virus aerosol propagation by positive airway pressure (PAP) circuit leak and mitigation with a ventilated patient hood. European Respiratory Journal 2020.

18. Tran K, Cimon K, Severn M, Pessoa-Silva CL, Conly J. Aerosol generating procedures and risk of transmission of acute respiratory infections to healthcare workers: a systematic review. PLoS One 2012; 7:e35797.

19. Bonilla N, Rojas MI, Cruz GNF, Hung S-H, Rohwer F, Barr JJ. Phage on tap–a quick and efficient protocol for the preparation of bacteriophage laboratory stocks. PeerJ 2016; 4:e2261.

20. Berlinski A. In vitro evaluation of positive expiratory pressure devices attached to nebulizers. Respiratory care 2014; 59:216–22.

21. Johnson DL, Lynch RA, Mead KR. Containment effectiveness of expedient patient isolation units. American journal of infection control 2009; 37:94–100.

22. Nishimura H, Fan Y, Sakata S. New applications of a portable isolation hood for use in several settings and as a clean hood. Journal of Thoracic Disease 2020; 12:3500.

23. Landry SA, Subedi D, MacDonald MI, et al. Point of emission air filtration enhances protection of healthcare workers against skin contamination with virus aerosol. Respirology 2022b.

24. Fanger PO, Christensen N. Perception of draught in ventilated spaces. Ergonomics 1986; 29:215–35.

25. Lednicky JA, Lauzard M, Fan ZH, et al. Viable SARS-CoV-2 in the air of a hospital room with COVID-19 patients. International journal of infectious diseases : IJID : official publication of the International Society for Infectious Diseases 2020; 100:476–82.

26. Leung NH, Chu DK, Shiu EY, et al. Respiratory virus shedding in exhaled breath and efficacy of face masks. Nature Medicine 2020:1–5.

27. Coleman KK, Tay DJW, Tan KS, et al. Viral load of severe acute respiratory syndrome coronavirus 2 (SARS-CoV-2) in respiratory aerosols emitted by patients with coronavirus disease 2019 (COVID-19) while breathing, talking, and singing. Clinical Infectious Diseases 2021.

28. Johnson G, Morawska L, Ristovski Z, et al. Modality of human expired aerosol size distributions. Journal of Aerosol Science 2011; 42:839–51.

29. Gaeckle NT, Lee J, Park Y, Kreykes G, Evans MD, Hogan Jr CJ. Aerosol generation from the respiratory tract with various modes of oxygen delivery. American journal of respiratory and critical care medicine 2020; 202:1115–24.

30. Mehta JH, Williams GW, I., Harvey BC, Grewal NK, George EE. The relationship between minute ventilation and end tidal CO2 in intubated and spontaneously breathing patients undergoing procedural sedation. PLOS ONE 2017; 12:e0180187.

31. McGain F, Bates S, Lee JH, et al. A prospective clinical evaluation of a patient isolation hood during the COVID-19 pandemic. Australian Critical Care 2021.

