## Supplementary for "Efficacy of a patient isolation hood in reducing exposure to airborne infectious virus in a simulated healthcare setting"

### Supplementary Materials

#### Modelling of physical aerosol data

The performance of the McMonty hood was assessed by comparing total particle number concentration PNC measured by the OPS in the room when the hood was active vs inactive. The effect was quantified by a fold reduction in the PNC observed for the two different operating conditions. To derive the fold reduction factor, we developed a simple particle number balance model charactering the rate of change in PNC expressed in Equation 1.

Equation 1

$$\frac{dPNC(t)}{dt}=\frac{E}{V}-\left( \frac{D}{100}\times PNC(t) \right)$$

Where *PNC* is time dependent total particle number concentration (#.^m-3^); *V* is the room volume (35.1 m^3^); *E* is the escape rate of particles (#.min^-1^) from the hood into room; and *D* is the rate of particle decay (#.min^-1^). Both, E and D, are assumed to be constant.

The first term on the right-hand side of the Equation 1 $\left( \frac{E}{V} \right)$ represents the rate of aerosol generation (source term) associated with particle escaping from the hood enclosure to the room. The second term $\left( \frac{D}{100}\times PNC(t) \right)$ represents the rate of particle decay or losses (sink term) caused by particle surface deposition and other removal mechanisms.

Equation 1 is an ordinary differential equation (ODE), and can be integrated with respect to time using the theorem given in Equations 2 and 3; solved for c using the boundary condition expressed in Equation 4; providing the final equation for PNC as a function of time in Equation 5.

Equation 2

$$y^{'}\left( t \right)=a.y\left( t \right)+b$$

Equation 3

$$y\left( t \right)=c.e^{at}-\frac{b}{a}$$

Equation 4

$$PNC\left( 0 \right)={PNC}_{i}$$

Equation 5

$$PNC(t)=\left( {PNC}_{i}-\frac{100E}{D.V} \right)e^{\frac{-D.t}{100}}+\frac{100E}{D.V}$$

Where PNC­_I_ is the initial particle number concentration in #.m^-3^.

PNC data were collected for two out of three experiment days (equipment was not present for one day due to COVID-19 travel conditions) with four nebulisation runs conducted each day. These PNC data were separated into two datasets for 1) McMonty active and 2) no containment conditions. One nebulisation run was excluded from the McMonty Active dataset as an equipment setup adjustment during testing led to additional particle escape, skewing results. This is shown in Supplementary Figure 1 for completeness.

**Supplementary Figure 1.** Plot of analyzed McMonty Active dataset and fitted curve alongside outlier run (blue data points) excluded from fit analysis.
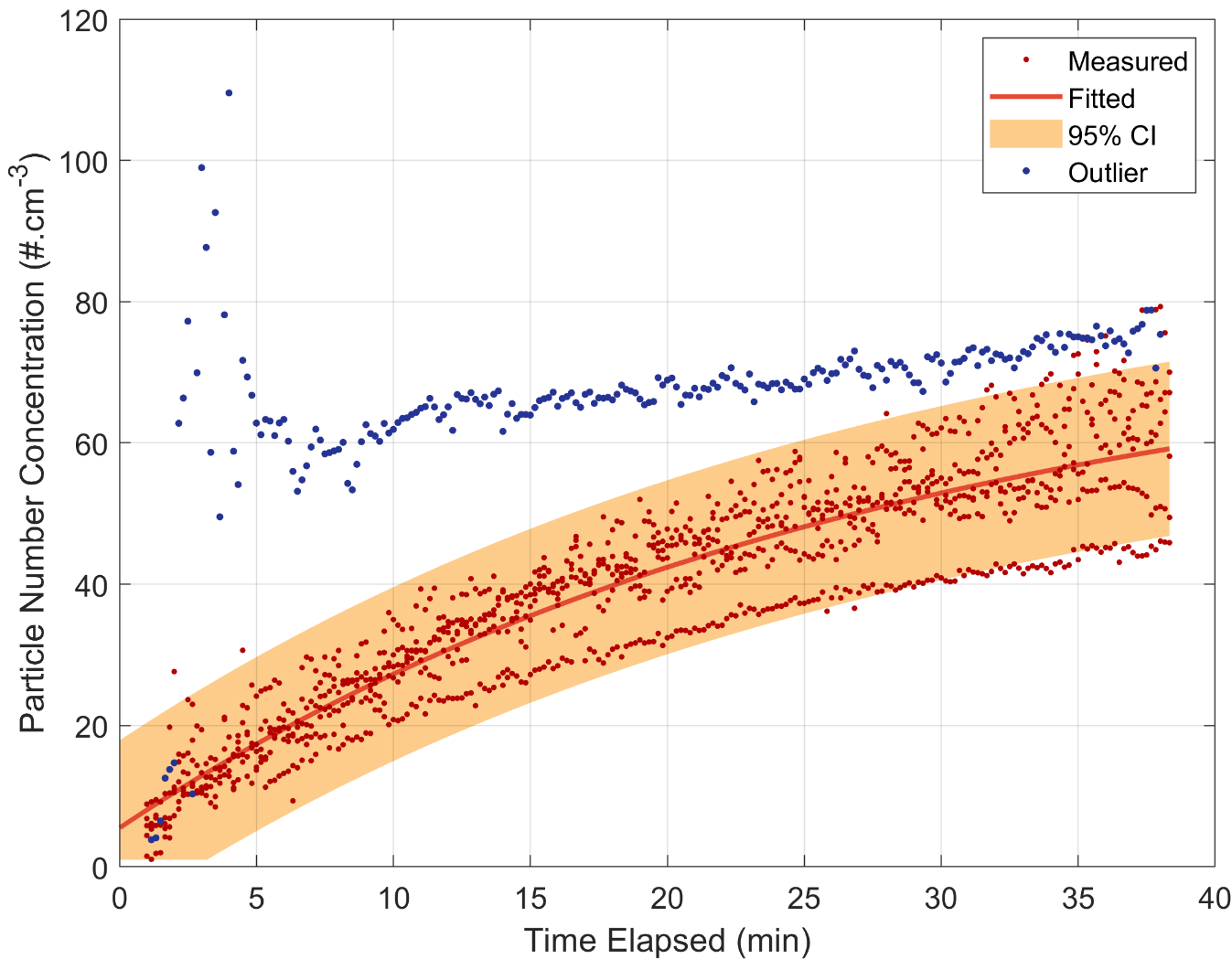

Datasets were truncated to remove all data before 60 seconds due remove the time delay between the nebuliser being switch on and a measureable PNC response being observed. Datasets were also truncated to remove all data after 2300 seconds to ensure the analysed datasets were of the same length. The model described through Equation 5 was fitted to these datasets in Matlab r2021b using a non-linear least squares fitting algorithm provided in the Curve Fitting Toolbox. The results of these fits is summarised in Supplementary Table 1.

To allow comparison with the virological analysis, an equivalent ‘Fold Reduction’ was calculated using Equation 6.

Equation 6

$$PF=\frac{E_{No\_Containment}}{E_{McMonty\_Active}}=\frac{(1.07 \pm0.03 ){\times10}^{10}}{(9.8 \pm0.4){\times10}^{7}}=109\pm5$$

Filtration efficiency is a related measure of aerosol protection equipment performance and is defined as the percentage of material stopped by the equipment. In this study the effective filtration efficiency (F) is calculated through Equation 7.

Equation 7

$$F=\left( 1-\frac{E_{McMonty\_Active}}{E_{No\_Containment}} \right)*100=\left( 1-\frac{\left( 9.8 \pm0.4 \right){\times10}^{7}}{\left( 1.07 \pm0.03 \right){\times10}^{10}} \right)*100=99.1\pm0.1$$

**Supplementary Table 1. Fitting results for the McMonty active and no containment PNC datasets.** Calculated terms are derived from fitting of particle number concentration data measured by optical particle sizer, ± standard deviation.

|  | **Escape Rate**  **(E)**  (#.min^-1^) | **Decay Rate**  **(D)**  (%.min^-1^) | **Adjusted r^2^** |
| --- | --- | --- | --- |
| **McMonty Active** | (9.8 ± 0.4) x 10^7^ | 3.6 ± 0.3 | 0.85 |
| **No Containment** | (1.07 ± 0.03 ) x 10^10^ | 4.2 ± 0.2 | 0.96 |

#### Recovery of viable bacteriophage following nebulisation using the BioSpot-VIVAS

The Biospot-VIVAS collects airborne particles (0.1-10 μm) using a laminar-flow water condensation method to maintain viral integrity and enable the recovery of infectious airborne virus in cell culture. To establish the feasibility of recovering viable PhiX174 from an air sample using the VIVAS, we first conducted tests using a custom fibreglass aerosol sampling chamber that could be operated within a class-2 biosafety cabinet (BSC2). Ten-fold dilution series of PhiX174 virus stocks were aerosolised into the sample chamber (290 mm x 600 mm x 600 mm, 81 L internal volume) using a commercial jet nebuliser (Aero-Neb Fortress Professional f1000. Aerogen, Galway, Ireland), and chamber air was collected using the BioSpot-VIVAS via a sample port. Each 30 minute sampling run consisted of 2x cycles of active nebulisation (airflow at 10 L/min) for 2 minutes followed by 13 minutes off, with continuous BioSpot-VIVAS sampling (full 30 mins). During experiment sampling periods, the following component temperature settings were employed: condenser at 5^o^C, initiator at 45^o^C, moderator at 12^o^C, sample platform at 10^o^C. These settings were maintained for later experiments in the simulated hospital room.

Nebulisation of phiX174 inoculum containing between 4.8 to 8.8 log_10_PFU was reliably recovered as viable sample using the BioSpot-VIVAS (Supplementary Figure 2). Below this inoculum range, recovery of phage became inconsistent (similar inputs would yield 0 – 3.5 log_10_PFU) and we did not successfully recover viable phage below nebulised inputs of 3.2 log_10_PFU in the aerosol chamber. Disinfection of the VIVAS collection column using 70% ethanol was sufficient to decontaminate up to 8.8 log_10_PFU of phage without affecting the collection efficiency of subsequent samples. Our results from the small aerosol chamber model validated a protocol for airborne phiX174 sampling with the BioSpot-VIVAS which we could apply to the simulated healthcare setting.

**Supplementary Figure 2.** **Scatterplot of total viable phage count (PFU) recovered in BioSpot-VIVAS sample (y-axis) compared to total phage count nebulised from inoculum (x-axis).** PhiX174 solution was completely aerosolised by medical jet nebuliser (airflow of 10 lpm) for 4 mins during 30 min sampling period. Total inoculum nebulised estimated from nebuliser optimisation at 1.5mL per 4mins. Blue line indicates linear regression trendline with 95% confidence interval (grey shading). Dashed line at lower limit of detection (1 PFU).

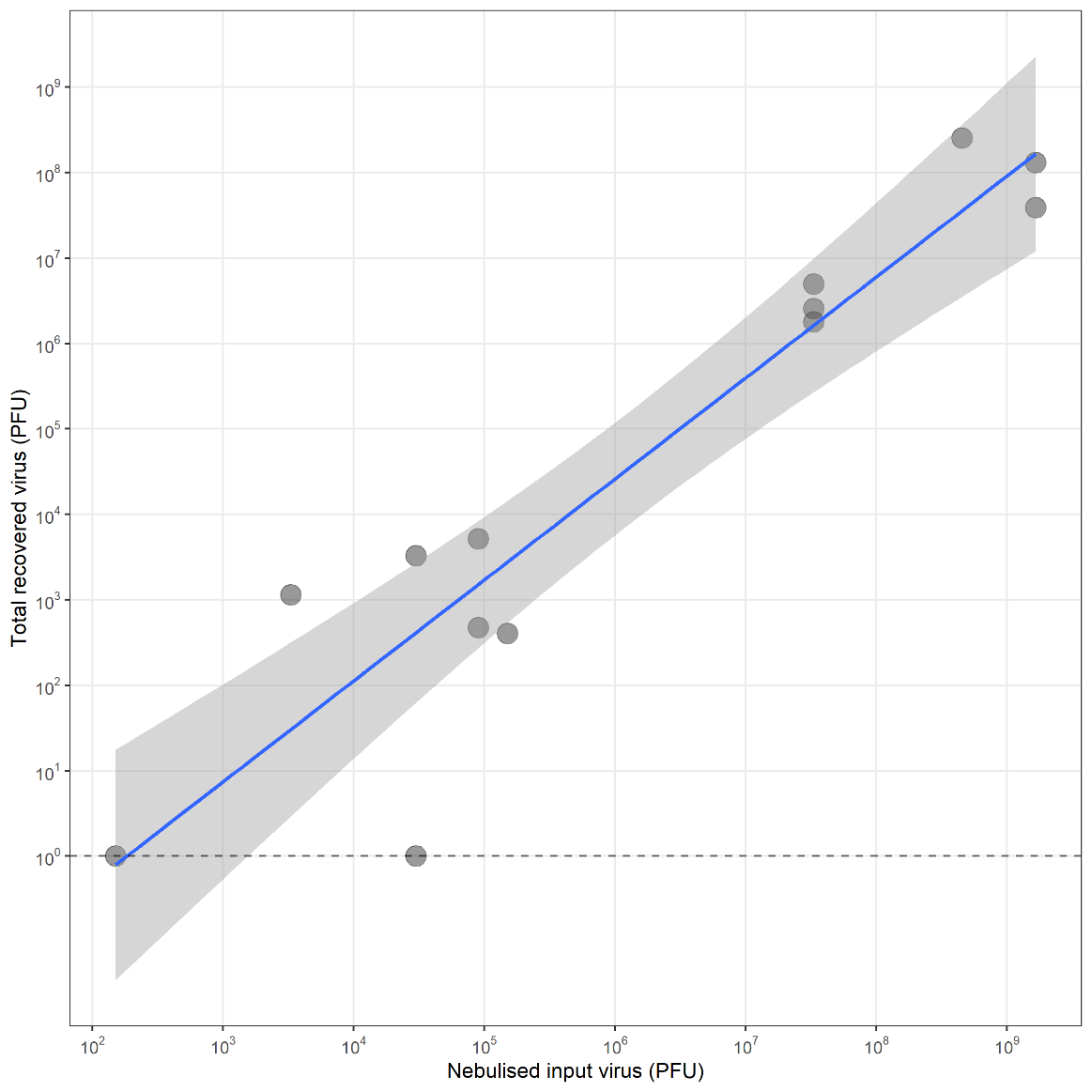

#### Dispersion range of nebulised bacteriophage in hospital simulation room

Based on previously published protocols used in the same facility by Landry et al. [1] we nebulised a 10mL solution of 10^8^ PhiX174 PFU/mL (for an effective total dose of 10^9^ viable phages) for ideal detection sensitivity by settle plates.

A new set of 90mm soft agar plates containing lawns of *E. coli* C bacterial host were positioned at 0.4-3.8m from the nebuliser for each aerosol generation experiment (13 plates per run; 10x flat, 3x hanging). Plates were exposed to the surrounding air during the 40 min nebulisation period, allowing the passive deposition of bacteriophage-laden airborne particles. Plates were then incubated overnight at 37 °C, and the number viral plaques was enumerated the following day, expressed as PFU per plate. In the control experiments, high phage counts (>100 PFU) were detected on a majority of settle plates when McMonty hood containment was inactive. We passively detected the presence of infectious airborne phage at distances up to 3.8m from the virus emission source (Supplementary Figure 3).

**Supplementary Figure 3.** **Scatterplot of total viable phage count (PFU) detected on settle plates (y-axis) relative to the distance (cm) of the settle plate to the nebuliser (x-axis).** Individual plaque counts for each settle plate when no containment was used during the sampling period (blue) compared to counts reached when active McMonty hood containment was used (red). Upper limit of detection per plate at 300 PFU (too many to count: TMTC).

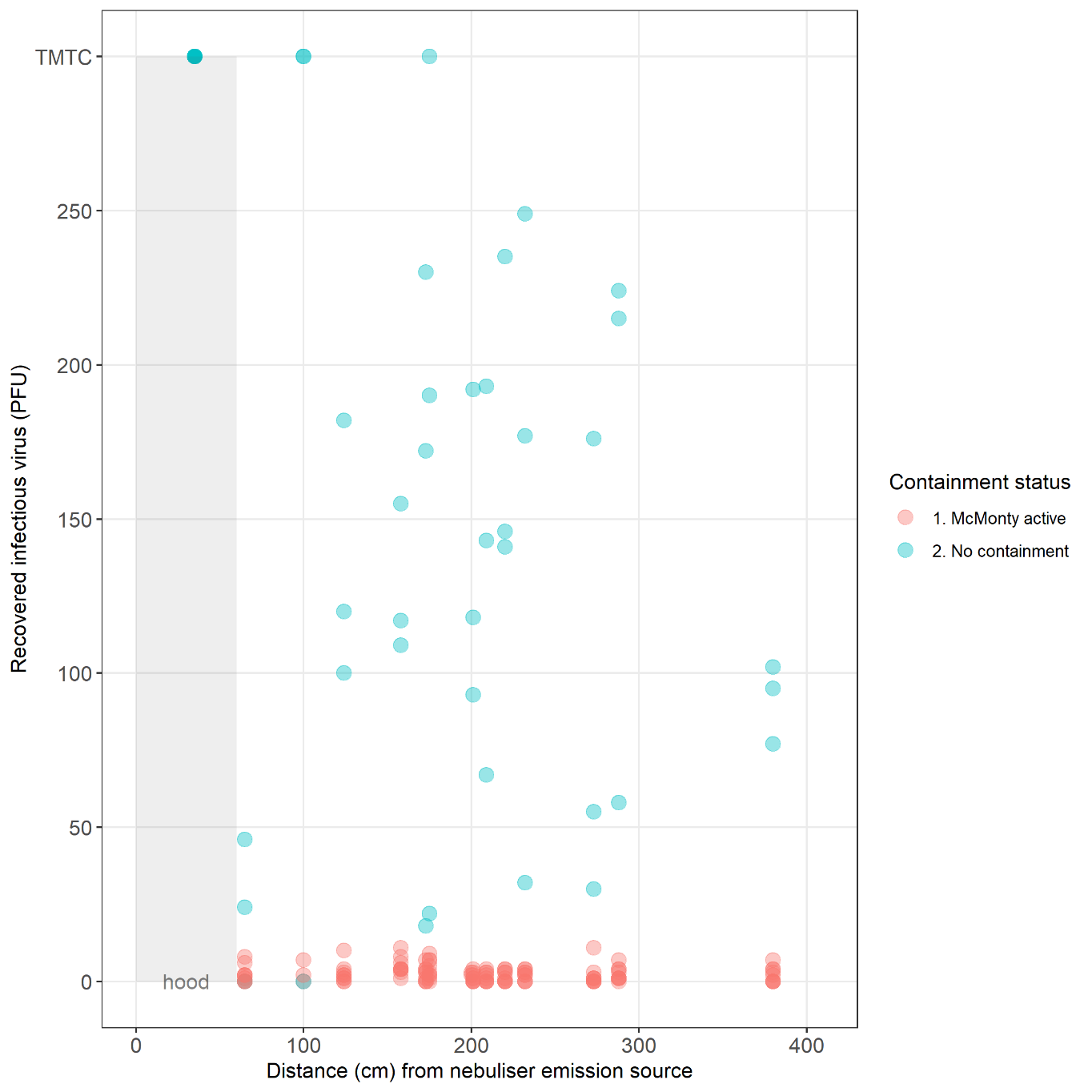

1.	Landry SA, Barr JJ, MacDonald MI, Subedi D, Mansfield D, Hamilton GS, Edwards BA, Joosten SA. Viable virus aerosol propagation by positive airway pressure (PAP) circuit leak and mitigation with a ventilated patient hood. *European Respiratory Journal* 2020.
